# Public’s perceived importance of non-pharmacological interventions for COVID-19 control in Greece: preliminary evidence from a cross-sectional study

**DOI:** 10.1101/2020.07.15.20153098

**Authors:** Eleni C. Boutsikari, Anna Christakou, Michail Elpidoforou, Ioannis Kopsidas, Nicholas Nikolovienis, Despina Kardara, Chrissoula C. Boutsikari, Christos Triantafyllou

## Abstract

**Background:** In the early stages of coronavirus disease 2019 (COVID-19) pandemic, while effective pharmaceutical approaches are pending, COVID-19 management relies primarily on non-pharmaceutical interventions (NPIs), such as social distancing, which requirepublic’s engagement and behavioral adjustment. This study aims to evaluate public’s perceived importance of the NPIs imposed for COVID-19 control in personal and public health protection in Greece.

**Methods:** This cross-sectional online study, enrolled 657 participants of the general Greek population in order to assess their beliefs and evaluate possible factors that influence their perceptions as regards NPI importance in personal and public health protection.

**Results:** Overall, Greeks considered NPIs important for health protection. The participants who were less likely to consider NPIs important were men (OR versus females=1.64, 95% CI:1.15 to 2.36, p=0.007), people younger than 40 years old (OR between ages over 40 versus ages below 40=0.48, 95% CI:0.34 to 0.68, p<0.001), and people who did not chose the Hellenic National Public Health Organization (EODY) to get informed about COVID-19 (OR of EODY versus other sources of information = 0.65, 95% CI:0.46-0.92, p= 0.014).

**Conclusions:** This study profiled Greek people who do and do not consider NPIs important, mainly as of their demographic features. Focused communicational strategies in certain population subgroups are recommended.

## Introduction

In December 2019, the first case of coronavirus disease 2019 (COVID-19) was reported in Wuhan China, signifying the beginning of a pandemic caused by SARS-CoV-2, a virus subsumed in a large family of zoonotic RNA single stranded viruses, known as corona viruses [1].Within three months SARS-Cov-2 spread globally and by March 31^st^2020, it had already caused 750,890 confirmed cases and 36,405 deaths worldwide [2]. The reproductive number (R0) of the virus was estimated close to 2.35, revealing its transmission potential in every area with at least four independently introduced cases [3].

In the early stages of COVID-19 pandemic, while effective pharmaceutical approaches are pending and environmental factors have proven insignificant in the epidemic control[4], COVID-19 management relies primarily on non-pharmaceutical interventions (NPIs), such as social distancing through permanent lockdown of institutional and professional facilities, and entertainment venues [5] or personal hygiene measures such as frequent hand wash [6]. Existing literature provides strong evidence that individuals’ perception of NPIs play a major role in successful disease mitigation, thus as NPIs currently constitute the centerline of COVID-19 control worldwide, research on public’s perception of NPIs is of extreme importance [7, 8].

COVID-19 research is already a vibrant field as a search for the term “COVID-19” on PubMed on May 5th, 2020 yielded more than 9,200 publications that include the term “COVID-19” in their title or abstract, a random search of which suggest that most, if not all of them, are indeed relevant to COVID-19. Despite the profound quick global reflexes on public health research, the vast majority of studies relevant to COVID-19 are of general medical[1] and pharmacological interest[9, 10], while less research is being done on the role of public’s perception in this global emergency.

### Objective

The aim of this study was to assess public’s beliefs and evaluate possible factors that influence public’s perceptions as regards NPI importance in personal and public health protection, in order to provide future recommendations that will enhance NPI effectiveness. The study was conducted in accordance with the recommendations of the Strengthening the Reporting of Observational studies in Epidemiology (STROBE) statement[11].

## Methods

### Study Design

This study was a cross-sectional online survey which started immediately after the national lockdown in Greece (March 23^rd^). We analyzed the preliminary data obtained from the participants between March 25^th^ and the first week of May 2020. Ethical approval was granted by the University of West Attica, Athens, Greece (ID:29341). Participants were asked to provide informed consent in order to gain access to the survey questions. All procedures were conducted in line with the Declaration of Helsinki [12].

### Setting and Recruitment

Respondents were recruited through Facebook, where research assistants and research coordinators distributed the survey to public Facebook groups. The online survey was hosted on “Qualtrics” [13], a secure data collection platform.

### Inclusion criteria

1. Participants are able and willing to provide informed consent
2. Participants are between 18 - 85 years old
3. Participants live permanently in Greece
4. Participants are Facebook users
5. Participants can read and write Greek

### Exclusion criteria

1. If participants live permanently outside Greece
2. If people cannot answer the survey questions and they do not understand Greek

### Data protection

This study was an anonymous online survey and participants were informed, before providing consent, that inclusion of personal information was not allowed. Data are kept in the platform and only study investigators sustain access to them, while no paper-based files were utilized.

### Survey Design

The study was based on the existing literature about perceptions of infectious disease control measures [7] and it was undertaken through a self-developed structured completion form, adapted to assess public’s perceptions as regards NPIs. The form consisted of 24 questions divided into 3 sections related to demographic factors, knowledge questions, beliefs about the importance of NPIs and self-reported adherence to home quarantine prior to national lockdown. Lastly, an open-ended field was provided for comments and feedback.

Certain epidemiological terms related to COVID-19 control were assessed for their comprehensibility in awareness and belief section, by using information included in the main guidelines for COVID-19 control, as provided by the Hellenic National Organization of Public Health (EODY)[14]. These terms were “incubation period” and “contact with SARS-CoV-2”, with the last being defined as one’s contact with a symptomatic confirmed COVID-19 case or an asymptomatic SARS-CoV-2 carrier, which is not followed by the individual’s symptomatic manifestation. Furthermore, as person in “high risk” for COVID-19 infection, was considered any respondent clinically compatible to COVID-19 or anyone who had travelled within March or had contacted another person clinically compatible to COVID-19, whereas “low risk” respondents were those who had not travelled, or met people with COVID-19 symptomatology or having manifested COVID-19 symptomatology themselves.

### Types of Bias

Biases associated with the verbal frame of surveys were avoided at the best applicable degree. Simple and comprehensible language was used, the valid answers in the questions with “right/wrong” design were the incorrect ones in order to minimize the halo effect[15], while optimal recall periods where adjusted in the domains of interest [16].

### Sample size calculation

In 2020, the Greek population was estimated in 10,430,130 people [17], with more than 80% of them sustaining Facebook accounts[18], thus Facebook users were considered a numerically representative source population. Sample size calculation based on the overall Greek population, with a confidence level of 95% and a 5% margin of error, determined that 385 respondents were required for the completion of the study, according to the Cochran’s formula [19].

### Variables

All questions apart from demographic factors were a posteriori scored in order be furtherly assessed. Section about knowledge and awareness of the NPIs included 10 multiple choice questions, with each one granted 1 point when answered correctly and 0 when answered incorrectly, thus the overall section score ranged from 0 to 10. In belief section 11 NPIs were given to be rated as of their importance for public and personal health, through horizontal 11-point numeric unipolar gliding scales ranging from 0 to 10, where 0 was “not important at all” and 10 “extremely important”, with the overall score of this section found between 0 and 110. Lastly, 4 questions relevant to self-reported adherence to home-quarantine were given, where every answer relevant to adherence was granted 1 point, and every opposite answer was granted 0 points, with the overall score of the section ranging from 0 to 4. All details about the scoring methodology are demonstrated on **Table 1**.

**Table 1:** Scoring system.

| Score definition | Minimum and Maximum Item Scores | Score Range | Mean, Median | Standard Deviation |
| --- | --- | --- | --- | --- |
| **Knowledge Score** |  |  |  |  |
| Row total of Item Scores in section of knowledge and awareness questions | 1 = every correct answer<br>0 = every incorrect answer | 1-10 | 7.06, 7 | 1.53 |
| **Perceived Importance Score** |  |  |  |  |
| Row total of item scores in section of belief questions (11 questions) | 10 = every NPI rated as extremely important<br>0 = every NPI rated as not important at all | 2-110 | 98.84, 100 | 16.22 |
| **Self-Reported Adherence to Home Quarantine** |  |  |  |  |
| Row total of item scores in section of self-reported adherence to home-quarantine (4 questions) | 1= every question relevant to:<br>a. Social contacts below normal<br>b. Home –quarantine> than 4 days/week<br>c. Voluntary adherence to home-quarantine according to the official guidelines/doctor’s advice<br>0= every question relevant to:<br>a. Social contacts above normal<br>b. Home-quarantine<4 days/week.<br>c. Non adherence to home quarantine, or adherence due to change of other people’s behavior or closure of entertainment venues | 0-4 | 2.59, 3 | 0.88 |

The numeric variables utilized to assess perceived importance of NPIs were the scores of knowledge and self-reported behavior as regards home quarantine. Demographic factors apart from sex and risk status, which naturally sustain two categories, were categorized into two subgroups. Since public’s perceptions were found different between post-secondary education and other education levels, as well as between areas with the most confirmed COVID-19 cases and the other areas of a country [7], post-secondary education was assessed separately from other education levels, likewise Athens from other areas of Greece. There is evidence that ages above 40 are related to higher possibility of infection [20] and higher case -fatality rate in Europe [21],thus age was categorized based on that cutoff point. As of cohabitation, people living with at least one infant or elder person were considered a different subgroup from those who did not. Formal government announcements were studied separately from every other source of information, in accordance to research approach of the existing literature [22].

### Data analysis

The analysis included only fully completed forms, with or without missing values. Missing values were addressed by the default approach of the statistical package which is the “listwise deletion” method [23]. Descriptive statistics and total responses were calculated for all variables and Spearman’s correlation coefficient was utilized for collinearity control among numeric variables. The outcome analyzed, namely the dependent variable, was the score regarding perceived importance of NPIs and it was dichotomized according to the “median split” method [24].Univariate analyses were performed to identify the exposure variables which demonstrate the greatest association with the outcome.

A multivariate logistic regression model was used to evaluate relationships between the outcome and the exposure variables, while odds ratios (OR) and their 95% confidence intervals (95%) were used to quantify these associations. Logistic regression was performed with the “forward stepwise” method, where the ratio of observations to the independent variables was kept above 5 [19]. Based on our study design, data were assumed to be independent and a p-value<0.05 was considered as statistically significant for all comparisons. The analysis was performed by the statistical package STATA ©, version 16.0.

## Results

### Demographic Data

The survey was initially completed by 669 individuals. Of those, 12 were excluded because they stated permanent residence outside of Greece. Demographic characteristics of the remaining 657 participants are described in **Table2**.

**Table 2:** Demographics of Study Participants.

| Variable |  | n | % | Total Responses |
| --- | --- | --- | --- | --- |
| **Sex** |  |  |  | **645/657** |
|  | Females | 227 | 35.19 |  |
|  | Males | 418 | 64.81 |  |
| **Age (years)** |  |  |  | **608/657** |
|  | 40 or above 40 | 330 | 50.23 |  |
|  | Below 40 | 327 | 49.77 |  |
| **Household members below 18 or above 70** |  |  |  | **657/657** |
|  | Yes | 279 | 42.72 |  |
|  | No | 378 | 57.53 |  |
| **Education level completed** |  |  |  | **642/657** |
|  | Under-graduate or Post-graduate education | 544 | 84.74 |  |
|  | Middle-High School or School | 98 | 15.26 |  |
| **Area of Permanent Residence** |  |  |  | **641/657** |
|  | Athens | 478 | 74.57 |  |
|  | Other | 163 | 25.43 |  |
| **Profession** |  |  |  | **644/657** |
|  | Healthcare | 187 | 29.04 |  |
|  | Not healthcare | 457 | 70.96 |  |
| **Risk status** |  |  |  | **644/657** |
|  | High | 44 | 6.83 |  |
|  | Low | 600 | 93.17 |  |
| **Source of Information** |  |  |  | **635/657** |
|  | EODY or formal government announcements | 348 | 54.80 |  |
|  | Other (doctor, greek or foreign medical associations or friend or COVID-19 positive person) | 287 | 45.20 |  |

### Knowledge of NPIs

Overall, the participants demonstrated high knowledge score (mean=7, Standard Deviation (SD) =1.53). Four hundred thirty-seven participants (67.96%) were aware of the term “contact with the new coronavirus”, as well as of all the main symptoms of COVID-19 (n=507, 78.97%). The term “incubation” though was understood only by 107 (16.56%) participants. Details about knowledge questions are provided in **Table 3**.

**Table 3:** Description of knowledge and awareness section.

| Questions | n | % | Score | Total Responses |
| --- | --- | --- | --- | --- |
| **Which are the main symptoms of COVID-19?** |  |  |  | **642/657** |
| Sore throat, dry cough , fever | 110 | 17.13 | 0 |  |
| Sore throat, dry cough , fever, fatigue, muscle and joint pain, dyspnea | 507 | 78.97 | 1 |  |
| Dry cough | 14 | 2.18 | 0 |  |
| None of the above | 11 | 1.71 | 0 |  |
| **What does come in “contact with COVID-19” mean?** |  |  |  | **643/657** |
| Having contacted a COVID-19 positive person, or an asymptomatic carrier and having no symptoms | 437 | 67.96 | 1 |  |
| Having contacted a COVID-19 positive person and having manifested symptoms | 66 | 10.26 | 0 |  |
| Not aware | 140 | 21.77 | 0 |  |
| **What does “incubation period” mean?** |  |  |  | **646/657** |
| The period from the first symptom to full recovery | 503 | 77.86 | 0 |  |
| The period from the infection to the first symptom | 107 | 16.56 | 1 |  |
| Not aware | 36 | 5.57 | 0 |  |
| **According to EODY the mean incubation period of SARS-CoV-2 is 5-6 days and the maximum is 14 days. If you contact a person who has the new coronavirus, how long should you stay at home-isolation?** |  |  |  | **643/657** |
| 5-6 days only upon symptom manifestation | 2 | 0.31 | 0 |  |
| 14 days regardless of symptom manifestation | 10 | 1.56 | 1 |  |
| 14 days only upon symptom manifestation | 628 | 97.67 | 0 |  |
| Not aware | 3 | 0.47 | 0 |  |
| **If I have respiratory symptoms, I should go to the closest hospital:** |  |  |  | **628/657** |
| Agree | 197 | 31.37 | 0 |  |
| Disagree | 431 | 68.63 | 1 |  |
| **If my respiratory symptoms get worse, I should go to the closest hospital:** |  |  |  | **636/657** |
| Agree | 546 | 85.85 | 0 |  |
| Disagree | 90 | 14.15 | 1 |  |
| **Flu vaccine protects from COVID-19:** |  |  |  | **611/657** |
| Agree | 11 | 1.80 | 0 |  |
| Disagree | 600 | 98.20 | 1 |  |
| **If I cough/sneeze on a tissue, hand washing is not mandatory:** |  |  |  | **609/657** |
| Agree | 18 | 2.96 | 0 |  |
| Disagree | 591 | 97.04 | 1 |  |
| **If I live alone, avoiding crowd is not necessary:** |  |  |  | **609/657** |
| Agree | 19 | 3.12 | 0 |  |
| Disagree | 590 | 96.88 | 1 |  |
| **It is safe, as regards COVID-19 transmission, to use personal objects of asymptomatic people?** |  |  |  | **612/657** |
| Agree | 23 | 3.76 | 0 |  |
| Disagree | 589 | 96.24 | 1 |  |

### Perceived Importance of NPIs

The mean score of the NPIs as of their importance in personal and public health protection was 98.84. The highest score in personal health protection was given to hand hygiene measure which demonstrated a mean score of 9.82 and the lowest score to mask and glove usage (mean = 6.73). In public health protection the same NPIs prevailed, with the highest importance score being given to hand hygiene and the lowest score to mask and glove usage, with mean scores 9.83 and 6.44, respectively.

### Self-reported behavior regarding social contacts

Self-reported adherence to home quarantine, was found above average, with a mean value equal to 2.59 (SD=0.88). Prior to the national lockdown, the majority of the participants (n=370, 58.92%) were capable of staying all day in the house from 0 to 4 days per week, while the remaining 258 (41.08%) of them could stay in the house from 5 to 7 days. The main barrier in one’s ability to apply home-quarantine proved to be work duties (n=317, 49.53%), and the main facilitator was the respondents’ sense of duty in adhering to NPIs (n=547, 86.41%). Detailed description about perceived importance of NPIs and self-reported adherence to social distancing are demonstrated in **Table 4**.

**Table 4:**
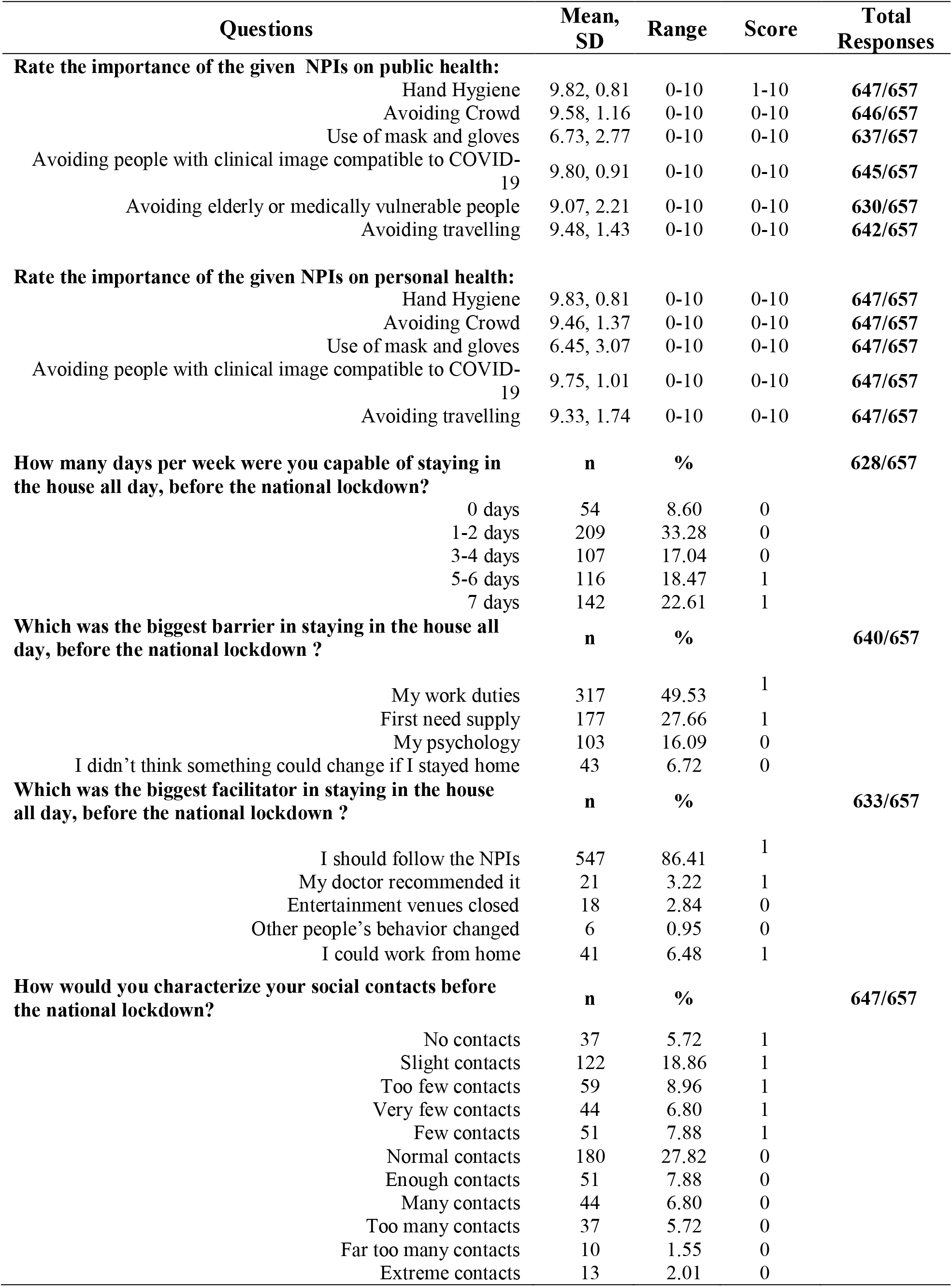
Description of beliefs and social contact section.

| Questions | Mean,<br>SD | Range | Score | Total<br>Responses |
| --- | --- | --- | --- | --- |
| **Rate the importance of the given NPIs on public health:** |  |  |  |  |
| Hand Hygiene | 9.82, 0.81 | 0-10 | 1-10 | **647/657** |
| Avoiding Crowd | 9.58, 1.16 | 0-10 | 0-10 | **646/657** |
| Use of mask and gloves | 6.73, 2.77 | 0-10 | 0-10 | **637/657** |
| Avoiding people with clinical image compatible to COVID-19 | 9.80, 0.91 | 0-10 | 0-10 | **645/657** |
| Avoiding elderly or medically vulnerable people | 9.07, 2.21 | 0-10 | 0-10 | **630/657** |
| Avoiding travelling | 9.48, 1.43 | 0-10 | 0-10 | **642/657** |
| **Rate the importance of the given NPIs on personal health:** |  |  |  |  |
| Hand Hygiene | 9.83, 0.81 | 0-10 | 0-10 | **647/657** |
| Avoiding Crowd | 9.46, 1.37 | 0-10 | 0-10 | **647/657** |
| Use of mask and gloves | 6.45, 3.07 | 0-10 | 0-10 | **647/657** |
| Avoiding people with clinical image compatible to COVID-19 | 9.75, 1.01 | 0-10 | 0-10 | **647/657** |
| Avoiding travelling | 9.33, 1.74 | 0-10 | 0-10 | **647/657** |
| **How many days per week were you capable of staying in the house all day, before the national lockdown?** | **n** | **%** |  | **628/657** |
| 0 days | 54 | 8.60 | 0 |  |
| 1-2 days | 209 | 33.28 | 0 |  |
| 3-4 days | 107 | 17.04 | 0 |  |
| 5-6 days | 116 | 18.47 | 1 |  |
| 7 days | 142 | 22.61 | 1 |  |
| **Which was the biggest barrier in staying in the house all day, before the national lockdown ?** | **n** | **%** |  | **640/657** |
| My work duties | 317 | 49.53 | 1 |  |
| First need supply | 177 | 27.66 | 1 |  |
| My psychology | 103 | 16.09 | 0 |  |
| I didn't think something could change if I stayed home | 43 | 6.72 | 0 |  |
| **Which was the biggest facilitator in staying in the house all day, before the national lockdown ?** | **n** | **%** |  | **633/657** |
| I should follow the NPIs | 547 | 86.41 | 1 |  |
| My doctor recommended it | 21 | 3.22 | 1 |  |
| Entertainment venues closed | 18 | 2.84 | 0 |  |
| Other people's behavior changed | 6 | 0.95 | 0 |  |
| I could work from home | 41 | 6.48 | 1 |  |
| **How would you characterize your social contacts before the national lockdown?** | **n** | **%** |  | **647/657** |
| No contacts | 37 | 5.72 | 1 |  |
| Slight contacts | 122 | 18.86 | 1 |  |
| Too few contacts | 59 | 8.96 | 1 |  |
| Very few contacts | 44 | 6.80 | 1 |  |
| Few contacts | 51 | 7.88 | 1 |  |
| Normal contacts | 180 | 27.82 | 0 |  |
| Enough contacts | 51 | 7.88 | 0 |  |
| Many contacts | 44 | 6.80 | 0 |  |
| Too many contacts | 37 | 5.72 | 0 |  |
| Far too many contacts | 10 | 1.55 | 0 |  |
| Extreme contacts | 13 | 2.01 | 0 |  |

### Multivariate Logistic Regression on Public’s Perceived Importance of NPIs

Spearman’s correlation coefficient was found low amongst numeric variables, namely 0.12 between knowledge and self-reported adherence to home-quarantine scores, indicating negligible collinearity [25], thus both variables were included in the analysis. P-value for the multivariate model was found statistically significant (p<0.0001).

According to multivariate logistic regression model respondents who did not consider NPIs important were more often found among males (OR versus females=1.64, 95% CI:1.15 to 2.36, p=0.007), among individuals younger than 40 years (OR between ages over 40 versus ages below 40=0.48, 95% CI:0.34 to 0.68, p<0.001), among participants residing permanently outside Athens (OR of Athens versus other areas=0.57, 95% CI:0.38 to 0.84, p=0.001) and among those who did not choose EODY to get informed about COVID-19 (OR of EODY versus other sources of information=0.65, 95% CI:0.46 to 0.92, p=0.014). Lastly, self-reported adherence to home-quarantine demonstrated a strong positive correlation with positive perceived beliefs of importance of NPIs, as for every answer relevant to home-quarantine adherence, there was a 23% multiplicative decrease in individuals’ perception of considering NPIs unimportant (OR=0.77, 95% CI:0.64 to 0.94, p=0.010). Regression analyses outcomes are demonstrated in **Table 5**.

**Table 5:**
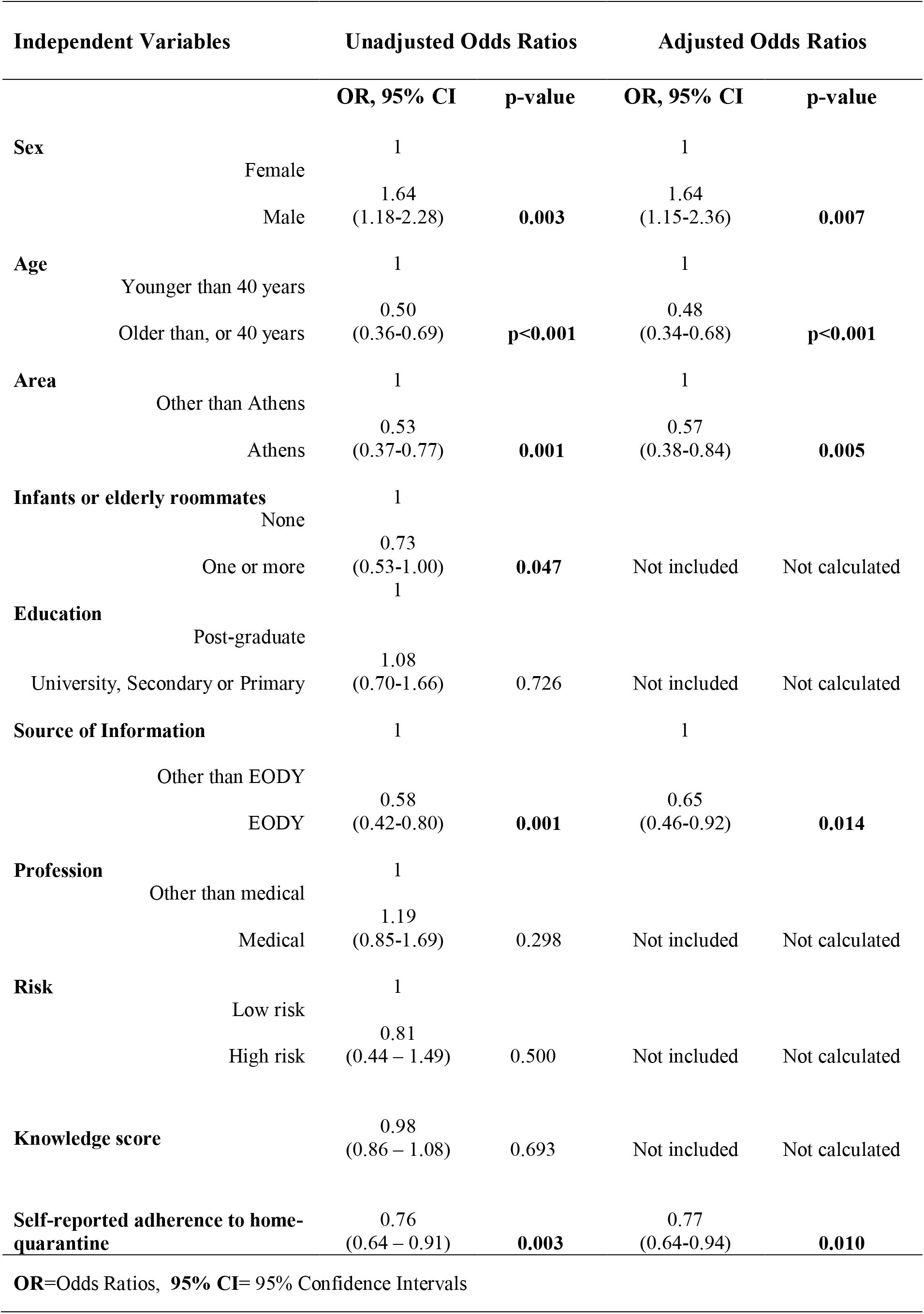
Univariate and multivariate regression analyses for factors that affect public’s perceptions about NPI importance in personal and public health protection (negative versus positive perceptions)

## Discussion

This research aimed to assess public’s beliefs and evaluate possible factors that influence public’s perceptions concerning importance of NPIs in health protection. A unique aspect of this study is that it estimates Greek population’s perceptions in the field of COVID-19, establishing the profile of the people who do not acknowledge the significance of NPIs in public and personal health. Apparently, a similar study has not been performed, thus we compared our findings with previous studies about attitudes and perceptions. This research correlated positively perceived importance of NPIs with self-reported adherence to home-quarantine, acknowledging thus the relation between beliefs and behavior, in agreement with established findings [26]. Another similarity was found for females who were more likely to consider NPIs important, held more positive attitudes [27], as well as better adherence levels to pandemic control measures [7]. Lastly, respondents from Athens, the area where the most COVID-19 confirmed cases were found, demonstrated higher perceived importance of NPIs, similarly to residents of Hubei when compared to people from other areas of China [7].

Differences were also found between this research and the existing literature. Conversely to previous evidence which did not correlate attitudes with age [27], in our study public’s positive perception of NPIs demonstrated strong correlation with ages greater than 40 years. Furthermore, the majority of the participants (n=348, 54.80%) chose EODY and formal government announcements as the most trusted source of information, a choice which led to positive perceptions about the significance of NPIs. The abovementioned result proves public’s trust and receptivity towards the formal authorities, an adversative to previous evidence finding, where the individuals were not influenced by formal communication strategies [28].

Greek community rated NPIs as important, a fact that possibly played a major role in Greece’s success in prompt COVID-19 control and early flattening of the epidemic curve [29].Although scoring high on knowledge questions and despite the respondents’ high education level, “incubation” appeared to be incomprehensible, since it was misinterpreted by the majority of the participants (77.86%) as the symptom period, a fact that consists an apparent knowledge gap. Future research should investigate the “knowledge gap hypothesis” in COVID-19, a phenomenon observed in other infectious diseases too [22], in which information is unequally distributed thorough a social system, where highly educated individuals are more properly informed than individuals of lower socioeconomic status. Provided that the sample of this study was mainly of high educational background, and yet unable to understand “incubation”, only assumptions can be made about knowledge gaps among people of different educational status.

Limitations of this study lie primarily on biases existing by the study design [15, 16] such as sampling bias, according to which the lack of internet access or social media profile excluded certain population subgroups from participation. Furthermore, this research is a cross-sectional study thus it can only provide a picture of a consecutive sample within a given timeframe. Neither validated scales nor instruments were utilized, thus measurement precision or errors cannot be estimated, despite the fact that numerous biases related to question forming were addressed. The sample demonstrated tendency towards male sex, a feature that presumably affected the outcome of regression analysis, as well as towards large urban centers as regards the area of permanent residence, post-secondary education and age below 70 years old. These features suggest that no conclusions can be drawn about population subgroups of different background and most importantly, about elderly people who are a medically vulnerable population and have been crucially hit in Europe by COVID-19 [21].

While this pandemic proceeds, health practitioners and government authorities worldwide are trying to profile COVID-19 through several approaches [4, 9]. Future research should primarily target on further investigation of public’s perception concerning the risk of COVID-19 transmission through asymptomatic carriers. Secondarily, the verbal frame of NPIs should be reassessed in order to be ensured that it is comprehensible and effective, while links between how knowledge gaps affect public’s behavior should be depicted. Lastly, communication promotion programs about COVID-19 mitigation should not only be carefully designed in order to achieve proper and equable information of the population, but targeted on the population subgroups that underestimate the importance of NPIs. The investigation of these issues will contribute in more effective and efficient management of prospective COVID-19 epidemic outbreaks and in enhancement of public health protection strategies.

## Data Availability

Dataset available upon request

## Contributors

EB conceived and funded the study. ME and IK contributed in the design of the form. CT and AC reviewed the form. All authors contributed in manuscript composition and revision before submission. The final draft was reviewed by CT.

## Acknowledgements

The authors gratefully acknowledge the support of the two anonymous reviewers for their guidance and the time they devoted to review this manuscript.

## Data Availability Statement

Dataset available upon request

## Compliance with Ethical Standards

### Research involving human participants and/or animals

This article does not contain any studies with animals performed by any of the authors. This article contains a cross-sectional online study with anonymous participation of humans.

### Statement of Human Rights

All procedures were performed in accordance with the ethical standards of the Institutional Research Committee which granted the ethical approval, the 1964 Declaration of Helsinki and its later amendments.

### Informed Consent

Informed consent was obtained from all individual participants included in the study, prior to participation.

### Patient consent for publication

Not required.

